# The genetic spectrum of febrile infection-related epilepsy syndrome (FIRES) and refractory status epilepticus

**DOI:** 10.1101/2023.02.12.23285754

**Authors:** Danielle deCampo, Julie Xian, Alexis Karlin, Katie R. Sullivan, Sarah M. Ruggiero, Peter D. Galer, Mark Ramos, Nicholas S. Abend, Alex Gonzalez, Ingo Helbig

**Author notes:** Corresponding author: Ingo Helbig, MD, Corresponding author’s address: Division of Neurology, Children’s Hospital of Philadelphia, Philadelphia, PA 19104, Corresponding author’s, Corresponding author’s. These authors contributed equally to this work and share first authorship (Danielle deCampo, Julie Xian, Alexis Karlin).

## Abstract

Febrile infection-related epilepsy syndrome (FIRES) is a severe childhood epilepsy with refractory status epilepticus after a typically mild febrile infection. The etiology of FIRES is largely unknown, and outcomes in most individuals with FIRES are poor. Here, we reviewed the current state-of-the art genetic testing strategies in individuals with FIRES. We performed a systematic computational analysis to identify individuals with FIRES and characterize the clinical landscape using the Electronic Medical Records (EMR). Among 25 individuals with a confirmed FIRES diagnosis over the last decade, we performed a comprehensive review of genetic testing and other diagnostic testing. Management included use of steroids and intravenous immunoglobulin (IVIG) in most individuals, followed by the ketogenic diet, and, after 2014, an increasing use of immunosuppressants, IVIG, and plasma exchange (PLEX). Genetic testing was performed on a clinical basis in almost all individuals and was non-diagnostic in all patients. We compared FIRES with both status epilepticus (SE) and refractory status epilepticus (RSE) as a broader comparison cohort and identified genetic causes in 36% of patients with RSE. In summary, despite the absence of any identifiable etiologies in FIRES, we performed an unbiased analysis of the clinical landscape, identifying a heterogeneous range of treatment strategies and characterized real-world clinical practice. FIRES remains one of the most enigmatic conditions in child neurology without any known etiologies to date despite significant efforts in the field, suggesting a clear need for further studies and novel diagnostic and treatment approaches. Furthermore, the difference in genetic signatures between FIRES and RSE suggest distinct underlying etiologies.

## Introduction

Febrile infection-related epilepsy (FIRES) is a rare and often catastrophic neurological condition characterized by refractory status epilepticus (RSE) that is preceded by a febrile illness occurring 2 weeks to 24 hours prior to onset of seizures [1]. Historically, FIRES was described in the pediatric population, but it is now recognized to occur in adults as well [2]. FIRES describes a subcategory of individuals with new onset refractory status epilepticus (NORSE), a clinical presentation characterized by de novo onset RSE that has no identifiable active structural, toxic, or metabolic cause [1]. While NORSE and FIRES represent a small fraction of all patients with RSE, they represent one of the most serious forms, with prolonged hospitalizations frequently followed by cognitive impairment and intractable epilepsy or death.

The underlying etiology for FIRES remains elusive. In some cases, an autoimmune or viral form of encephalitis is identified, although in most cases no underlying cause is identified. These cases are designated as cryptogenic and are an area of active investigation [1]. A fulminant aberrant inflammatory response in the central nervous system has been proposed as a unifying mechanism [3, 4]. SE may induce a proinflammatory cascade, with several of these molecules promoting proconvulsant activity [5, 6]. Furthermore, reported abnormal imaging findings have been variable and nonspecific [7, 8].

Among studies focused on elucidating the etiology of FIRES, there has been an increased focus on potential underlying genetic factors as causative etiologies, though no genetic factors have ever been definitively identified. The lack of genetic explanation in FIRES stands in stark contrast to the genetic landscape of epilepsy more broadly in which the genetic yield is up to 33% with different forms of genetic testing [9, 10]. Studies exploring genetic etiologies of SE and RSE indicate a subset of various genes that are associated with SE [11, 12]. However, there are fewer recognized genes and validated variants associated with RSE [13, 14].

Given the rarity of NORSE and FIRES, a consistent barrier to identifying genetic etiologies is the limited population from which data can be systematically analyzed. While there have been reports of individuals with FIRES or NORSE having variants in genes associated fever-sensitive epilepsies or metabolic diseases,[15, 16, 17, 18, 19] many of these variants have not been confirmed to be explanatory, further contributing to the air of enigma regarding FIRES. A genetic susceptibility to immune dysregulation through variants in the cytokine pathway has also been suggested [15] but systematic genetic studies have not replicated these findings [20, 21]. Recently, we published exome findings on 50 individuals with FIRES and did not identify any disease-causing genetic variants, including in candidate genes. Human leukocyte antigen (HLA) sequencing in a previous cohort of 29 individuals with FIRES previously failed to identify prominent HLA genes [21]. Consequently, the role of genetics as an etiology or predisposition for FIRES remains inconclusive.

Therefore, in this retrospective study, we aimed to further delineate the genetic etiology of FIRES, as well as SE and RSE more generally. We used natural language processing (NLP) methods to identify children diagnosed with FIRES at a large tertiary center and systematically reviewed the genetic testing completed as part of their clinical care to identify a potential genetic etiology for FIRES. To determine how the genetic yield of FIRES fits within the broader landscape of children with non-FIRES related SE and RSE, we examined broader cohorts of children diagnosed with SE or RSE as their first presentation of seizure.

## Methods

### Identification and inclusion of individuals with FIRES

This was a retrospective single-center observational study performed at Children’s Hospital of Philadelphia (CHOP). Individuals with FIRES were identified through NLP of free-text patient notes from the Electronic Medical Records (EMR) between 2013 and 2023 using the search terms “Febrile infection-related epilepsy” or “FIRES” or “new-onset refractory status epilepticus” or “NORSE” with “febrile infection.” Some individuals in our study were initially admitted to outside institutions prior to transfer to CHOP as a tertiary care center. We included these patients as we had access to shared EMRs from their hospitalizations at the time of their subsequent presentations to CHOP. We manually reviewed the selected charts to confirm the diagnosis of FIRES based on formal consensus criteria [1].

The following definitions were used. Status epilepticus (SE) was defined as 5 minutes or more of continuous clinical seizure activity or recurrent seizure activity without recovery between seizures. Refractory status epilepticus (RSE) was defined as SE persisting despite administration of at least two appropriate parenteral medications including a benzodiazepine, without a specific duration required. We used consensus guideline definitions of NORSE and FIRES [1]. NORSE was defined as *de novo* onset of RSE without an identifiable acute or active structural, toxic, or metabolic cause. FIRES was considered a subcategory of NORSE requiring prior febrile illness starting between 2 weeks and 24 hours before onset of RSE (with or without fever at onset of status epilepticus). Individuals were excluded if: (1) there was no documentation of preceding illness, remote history of illness without documentation, or lack of outside records to confirm preceding febrile illness; (2) the word “FIRES” was included in the chart but referred to family history or to a publication that included the search term or the differential diagnosis where FIRES was ultimately dismissed; or (3) the clinical history did not align with the formal consensus definition for FIRES. Additionally, three individuals with NORSE but without sufficient clinical information to diagnose FIRES based on the characteristics defining FIRES described above were excluded. This study was completed and approved by the Institutional Review Board at CHOP.

### Clinical and treatment data

We manually reviewed demographic and clinical data of 25 individuals with FIRES, including information on hospital admissions and genetic workup (see below). For each patient, we reviewed their prior medical and developmental history, history of preceding illness with fever, and age at time of hospitalization for FIRES and at discharge. For treatment strategies, all children were placed on anesthetic infusions during their hospital course (midazolam, pentobarbital, ketamine); however, detailed records about the dates of administration were limited so we focused only on whether anesthetic infusions were administered, number of anti-seizure medications at discharge, and the use of use of anti-inflammatory and immunomodulatory agents including intravenous immunoglobulins (IVIG), steroids, plasmapheresis (PLEX), and other immunomodulatory agents.

### Genetic testing

For individuals with confirmed diagnoses of FIRES, a comprehensive review of patient records was performed to identify elements of their diagnostic workup for FIRES, with particular attention to the timing and type of genetic testing. Previous genetic testing was identified in the EMR and stored as laboratory results, external media uploads, or within recent neurology and genetic clinical notes. Lastly, a free text search of medical charts was performed with the key words: “*SCN2A*”, “*POLG*”, “gene”, “karyotype”, “microarray”, “epilepsy panel”, “exome”, and “mito”. Cases where genetic testing was performed but reports were not available for review were noted. We documented the type of genetic test (single gene sequencing, karyotype, microarray, whole exome sequencing, mitochondrial genome sequencing), results, report date, coding and protein variant change, inheritance, and variant pathogenicity.

### Cohorts for mapping the genetic etiologies of SE and RSE more broadly

To characterize the clinical and genetic landscape of individuals with SE and RSE more broadly we used two cohorts: (1) 32,112 individuals with epilepsy in the Pediatric Epilepsy Learning Health System (PELHS) and (2) 1,894 individuals with presumed or confirmed genetic epilepsies at CHOP. First, using the PELHS cohort, we mapped the genetic and clinical landscape of non-FIRES RSE through systematic evaluation of the EMR, capturing more than 4.5 million full-text health care notes spanning 203,369 total patient-years. We limited our search in this cohort to the first presentation of RSE, as subsequent encounters with RSE documented in the patient notes could refer to a new RSE event or history of prior RSE. We assessed the overall distribution of RSE onset in this larger cohort and contrasted it with the wider distribution of onset in FIRES.

Secondly, in order to generate an understanding of genes that may be implicated in SE and RSE more broadly, we further delineated the genetics of children with SE and RSE using a dataset comprised of 1,894 individuals with presumed genetic epilepsy and neurodevelopmental disorders and characterized the genetic yield of SE. This resulted in 1,158 individuals in the CHOP cohort with SE. When performing NLP on patient notes to capture phenotypes such as the presence of certain seizure types of neurological features, we only parsed notes prior to a genetic diagnosis if applicable. The rationale was to control for bias associated with clinical impressions following a molecular diagnosis. For example, individuals with *PCDH19*-related disorders might have ‘status epilepticus’ in their patient charts due to a general clinical description of *PCDH19*-related disorder, rather than the patient’s specific phenotype.

In our cohort of 1,894 individuals with presumed genetic epilepsies, we then focused only on individuals with a known genetic diagnosis who presented with RSE at first seizure presentation (i.e., no prior history of epilepsy) in order to identify genetic etiologies associated with a NORSE-like presentation. We identified 208 individuals with RSE from NLP. Seventy-five of these individuals had a genetic diagnosis. Twenty-two of the 75 individuals presented with RSE at their first seizure presentation. We then manually reviewed patient charts of this smaller cohort and excluded the following: (1) individuals who did not have true RSE (e.g., in whom “refractory” referred to seizure action plans embedded in the chart rather than the clinical history); (2) individuals who had a pre-existing diagnosis of epilepsy; (3) one individual in whom the genetic diagnosis did not correlate with the RSE presentation; and (4) neonatal-onset RSE.

We focused on genetic etiologies associated with RSE after the neonatal period to serve as a closer comparison group to FIRES/NORSE. This left us with four individuals carrying three separate genetic diagnoses. This subgroup also included two additional individuals. The first was the twin of another individual, carried the same clinical presentation (4 months earlier), and was diagnosed with the same genetic condition. The second had been previously excluded from our FIRES database as she did not meet FIRES diagnostic criteria. This left six individuals who had presented with RSE as first seizure presentation and who were ultimately diagnosed with a genetic epilepsy syndrome.

Lastly, to better understand whether children with first onset of seizure presentation of SE and in a subgroup of individuals with RSE who had a genetic diagnosis had a different clinical phenotype than those without a genetic diagnosis, we analyzed phenotypic features in individuals with SE stratified by individuals with and without an identified genetic etiology, calculating odds ratios with 95% confidence intervals using Fisher’s exact test. Phenotypic associations between the two subgroups are presented as a phenogram, a previously published method that allows us to visualize the overall constellation of a selection of phenotypic features and severity of clinical presentations between groups [22, 23, 24].

## Results

### Individuals with FIRES can be identified from the EMR in a large tertiary health network

Using NLP methods, we identified 201 individuals. We then filtered out individuals where FIRES was used in: (1) the context of a publication reference that contained the word “FIRES” or for conditions that have been documented to be associated with FIRES or NORSE, or (2) patients for which a referral note was written but the individual was never evaluated at CHOP. This resulted in 59 individuals for manual chart review. Thirty-four individuals were excluded due to an alternative diagnosis, not meeting criteria for FIRES, or a remote history of FIRES with absence of documentation of the illness. There were eight individuals diagnosed with FIRES but excluded because of a lack of clinical information (Supplementary Table 1). There were six patients with a presumed diagnosis of NORSE, but only three had sufficient clinical documentation to confirm diagnosis. One of these patients was also diagnosed with MOG encephalitis. Notably, one patient had a diagnosis of *PCDH19*; however, for this individual there was insufficient documentation for further study (Supplementary Table 2). Following exclusion after manual review, there were 25 individuals diagnosed with FIRES with sufficient clinical histories (Figure 1).

**Figure 1.**
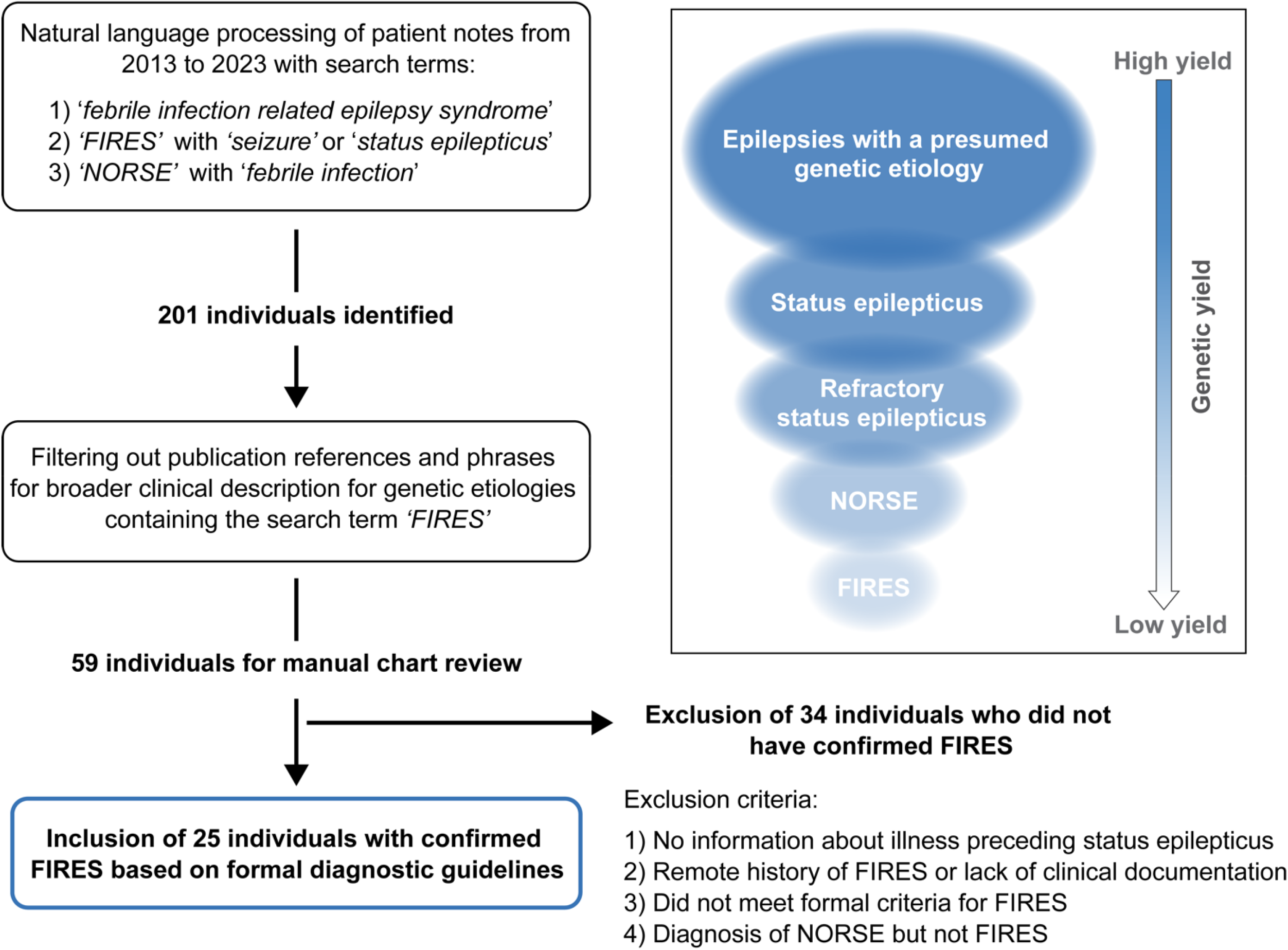
Framework for the identification of individuals with Febrile Infection Related Epilepsy Syndrome (FIRES). Natural language processing was applied across a large pediatric institution Electronic Medical Record database followed by manual chart review, leading to identification of 25 individuals with confirmed FIRES based on formal consensus criteria. FIRES is a subset of New Onset Refractory Status Epilepticus (NORSE), showing the hierarchy of FIRES and NORSE with broader subgroups of individuals with SE related epilepsies and epilepsies with a presumed underlying genetic cause (inset).

In the 25 individuals with FIRES, the median age of onset of SE was 7.25 years (IQR 5.06 – 9.84 years) (Figure 2A). Eleven individuals (44%) were female. Twenty individuals (80%) had no significant prior medical history. Of the five individuals with a prior medical history, one individual had a history of two simple febrile seizures that occurred more than one year prior, as well as recurrent urinary tract infections. Other medical conditions included autism, attention deficit hyperactivity disorder, concussion diagnosed two weeks prior to seizure onset, and celiac disease (Table 1). Twenty-four (96%) were developmentally typical prior to seizure onset. Eight individuals (32%) had fever and fatigue as the preceding symptoms prior to seizure onset, while eight individuals (32%) had headaches, with two of these individuals also presenting with altered mental status. Five individuals (20%) had gastrointestinal symptoms of vomiting and/or diarrhea, and one individual also had a rash. Four individuals (16%) had upper respiratory infection prodromal symptoms. Five individuals had an identified bacterial or virus (*rhinovirus*/*adenovirus*/*Bartonella, Mycoplasma, Metapneumovirus, rhinovirus*/*parvovirus, E. coli* urinary tract infection). Three individuals were ultimately diagnosed with autoimmune encephalopathy with anti-thyroid encephalitis (anti-thyroid peroxidase antibodies), Sjogren’s syndrome, and catastrophic antiphospholipid syndrome.

**Figure 2.**
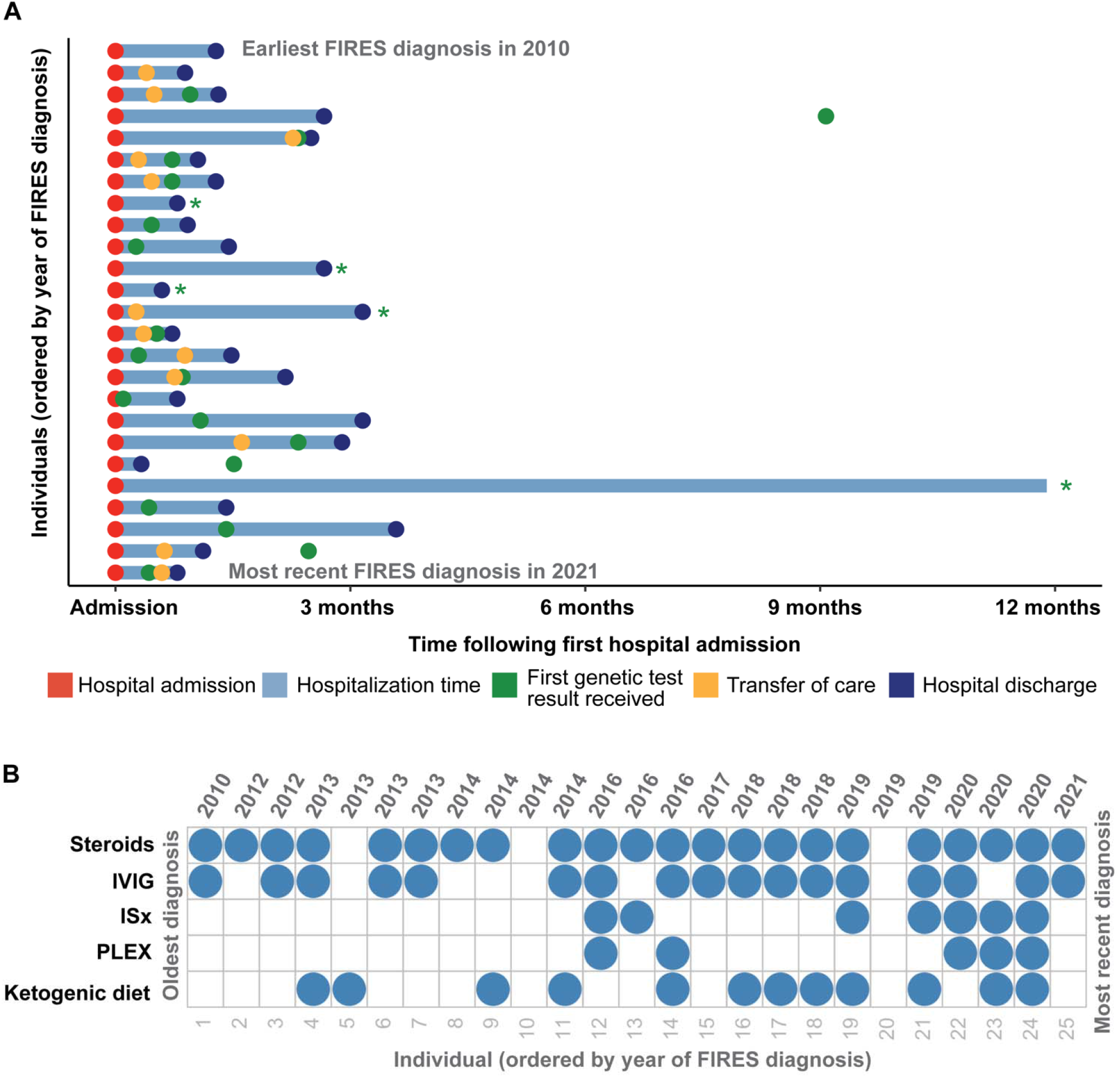
Clinical histories of 25 individuals with FIRES identified in the Electronic Medical Records (EMR), highlighting in **(A)** time at hospital admission and discharge in addition to transfer of care and time of first genetic test results received in the first year following first status epilepticus event. Individuals whose genetic testing results were received more than a year after first hospital admission are indicated with a green asterisk. **(B)** Overview of treatment strategies for each individual presenting with FIRES, ordered by year of FIRES diagnosis, showing common use of steroids and intravenous immunoglobulins (IVIG) and an increased use of immunosuppressants (ISx) and plasmapheresis (PLEX) after 2014.

**Figure 3.**
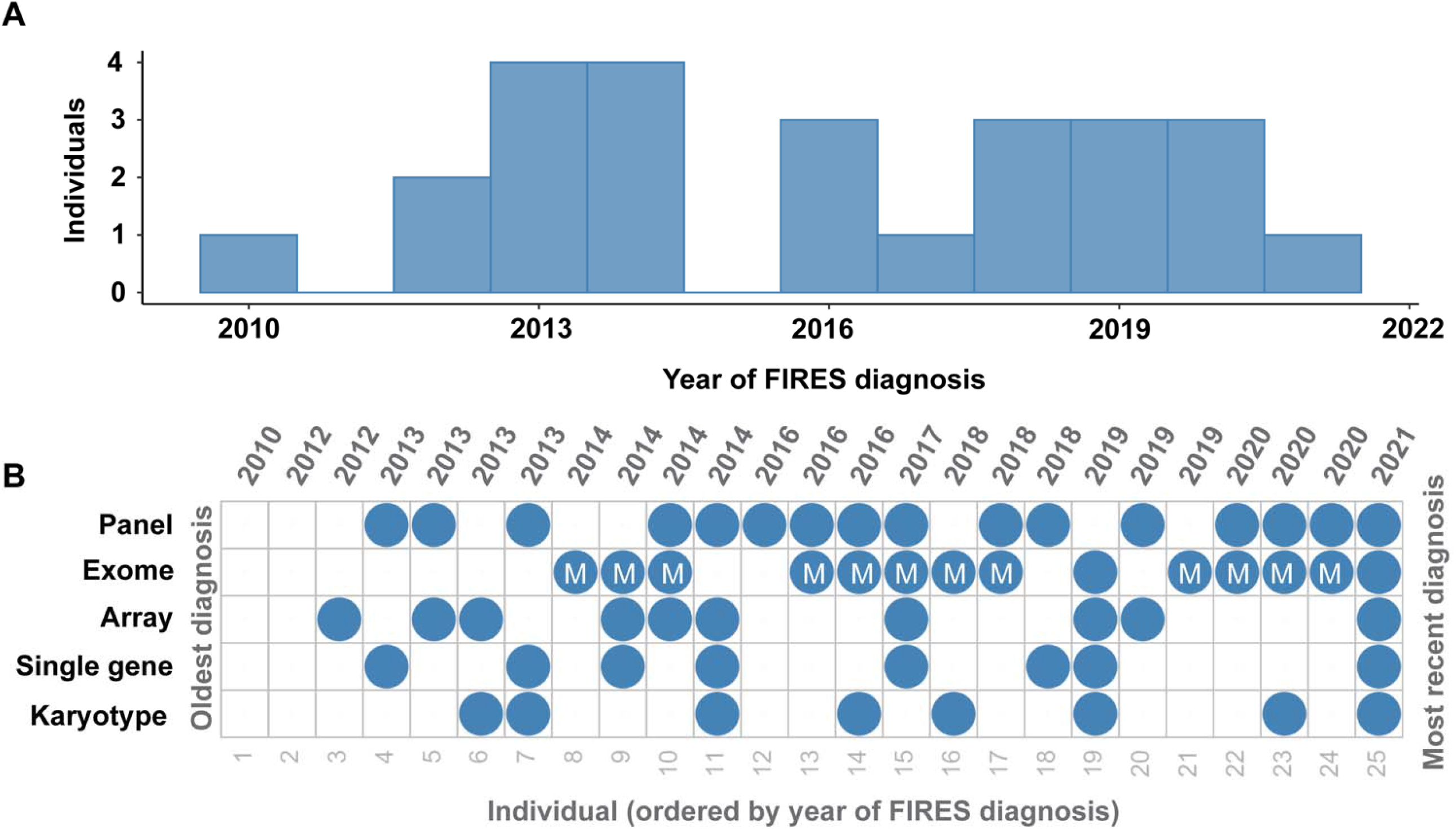
Genetic testing in FIRES has been heterogeneous over the years. **(A)** Distribution of FIRES diagnoses in the past decade. **(B)** Overview of genetic testing including karyotype, single gene testing, microarray, whole exome sequencing (with mitochondrial testing indicated), and gene panel in 25 individuals with FIRES, ordered from the oldest diagnosis in 2010 to most recent diagnosis in 2021.

The median duration of hospitalization before discharge or transfer of care was 1.3 months (IQR 27-81 days). One individual remained hospitalized due to social factors and had been in the hospital for a duration of 2.8 years up to the time of inclusion. All children were noted to have some form of cognitive, speech, or physical impairment at the time of discharges (Table 1). Six (24%) individuals died before hospital discharge. One individual that was diagnosed with FIRES died seven years later, due to cardiac arrest secondary to a variant in *SCN5A*.

**Table 1.** Demographics and clinical histories of 25 individuals with FIRES.

| Participant | Year diagnosed with FIRES (age, years)**** | Sex | Prior medical diagnoses | Development | Prodromal symptoms* | Positive findings** | Hospital duration (months) | Death before discharge |
| --- | --- | --- | --- | --- | --- | --- | --- | --- |
| 1 | 2010 (5-10 years) | F | none | normal | Headache |  | 1.28 | N |
| 2 | 2012 (5-10 years) | M | none | normal | URI |  | 0.89 | N |
| 3 | 2012 (5-10 years) | M | none | normal | Headache, Altered mental status |  | 1.32 | Y |
| 4 | 2013 (10-15 years) | M | concussion 2 weeks prior | normal | Headache, Altered mental status |  | 1.28 | Y |
| 5 | 2013 (5-10 years) | M | none | normal | URI |  | 2.50 | N |
| 6 | 2013 (15-20 years) | F | none | normal | none | Mycoplasma IgM/IgG+ | 2.66 | N |
| 7 | 2013 (10-15 years) | M | none | normal | Headache |  | 1.05 | N |
| 8 | 2014 (0-5 years) | F | none | normal | GI | Metapneumovirus;SCN5A | 2.66 | N*** |
| 9 | 2014 (5-10 years) | M | ADHD | normal | URI | +Rhino/parvovirus | 1.44 | N |
| 10 | 2014 (0-5 years) | F | none | normal | Fatigue |  | 0.79 | N |
| 11 | 2014 (5-10 years) | F | none | normal | Headache |  | 0.92 | Y |
| 12 | 2016 (0-5 years) | M | none | normal | Fatigue |  | 3.16 | N |
| 13 | 2016 (10-15 years) | F | none | normal | None | Sjogren's syndrome | 0.59 | N |
| 14 | 2016 (5-10 years) | M | none | normal | Headache |  | 0.72 | Y |
| 15 | 2017 (0-5 years) | F | 2 febrile seizures, recurrent UTI | normal | Fatigue | E. coli+ UTI | 1.48 | N |
| 16 | 2018 (5-10 years) | M | none | normal | URI/Sinusitis |  | 2.17 | N |
| 17 | 2018 (0-5 years) | M | none | normal | GI |  | 3.16 | N |
| 18 | 2018 (5-10 years) | F | none | normal | Headache |  | 0.79 | N |
| 19 | 2019 (5-10 years) | M | autism | autism | None |  | 34.3 | N |
| 20 | 2019 (0-5 years) | M | none | normal | GI | Adeno/Rhinovirus, bartonella | 0.33 | N |
| 21 | 2019 (5-10 years) | M | none | normal | Headache |  | 2.89 | Y |
| 22 | 2020 (15-20 years) | F | Celiac disease | normal | GI | lupus cerebritis/ catastrophic APS; +GAD | 1.11 | Y |
| 23 | 2020 (5-10 years) | F | none | normal | GI, Rash |  | 3.58 | N |
| 24 | 2020 (5-10 years) | F | none | normal | None | Anti-TPO | 1.41 | N |
| 25 | 2021 (15-20 years) | M | none | normal | None |  | 0.79 | N |

\*All patients had fever per consensus guidelines for FIRES; \*\* determined to be not causative for FIRES; \*\*\*death occurred 7 years after discharge
\*\*\*\* Ages are provided in ranges of 5 years.
ADHD attention deficit/hyperactivity disorder, UTI urinary tract infection, URI upper respiratory infection, GI gastrointestinal symptoms, TPO thyroid peroxidase, IVIG, PLEX, steroids, immunomodulatory

### Treatment in FIRES has changed across the years

All individuals were placed on anesthetic infusions for treatment of seizures during their hospitalization. Of the 19 individuals discharged, all individuals were on anti-seizure medications (ASMs) with a median of three ASMs (IQR 3–4). Specific ASMs are listed in Table 2. Twenty-two (88%) individuals received some form of immunotherapy (IVIG, steroids, PLEX, immunomodulatory agents). Twenty-two (88%) received steroids, seventeen (68%) received IVIG, seven (28%) received immunomodulatory agents (included anakinra, tocilizumab, rituximab, cyclophosphamide and hydrochloroquine), and five (20%) received PLEX (Figure 2A). Twelve (48%) individuals were initiated on the ketogenic diet during their hospitalization. While there was a heterogeneous pattern of treatment strategies over time (Figure 2B), we found that immune-mediated strategies increased after 2014 with increased use of ketogenic diet and immunomodulating therapies.

**Table 2.**
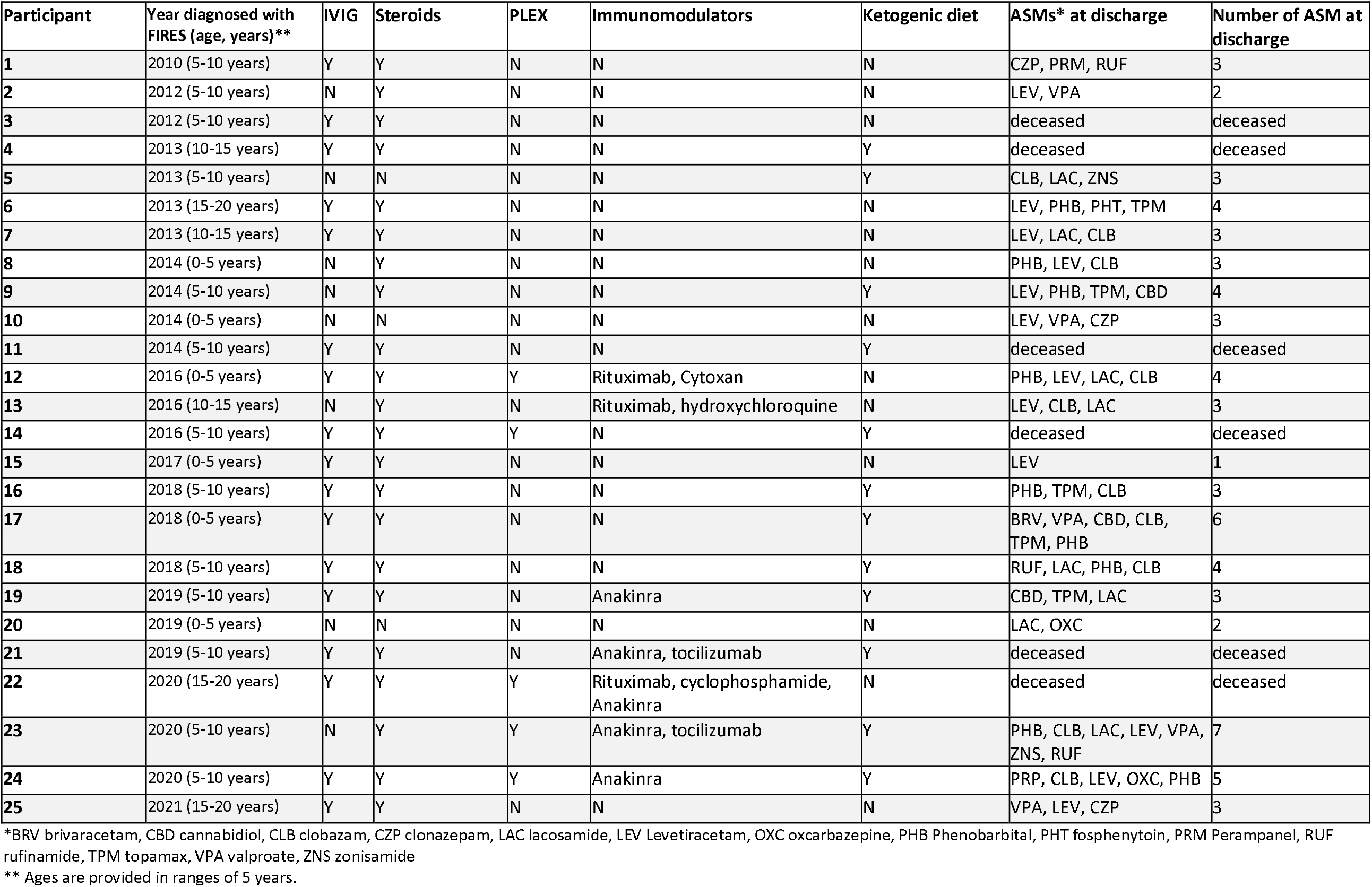
Treatment strategies in FIRES.

### Genetic testing is heterogeneous in FIRES and does not reveal underlying genetic etiology

Twenty-three (92%) individuals received genetic testing. Fifteen individuals received the results of testing during their hospital admission, and eight received results of genetic testing after discharge. Among the 23 with genetic testing, eight (35%) received single gene testing, eight (35%) received karyotypes, twelve (52%) received SNP chromosomal microarrays (two of whom did not have additional testing), eighteen (78%) received epilepsy panels, thirteen (57%) received exomes, and eleven (48%) received mitochondrial sequencing. One individual also received Fragile X testing, and three individuals had an exome reanalysis performed. No individuals in our cohort received whole genome sequencing, although some families were offered this testing (Supplementary Table 3).

Systematic review of genetic testing revealed no genetic etiology for FIRES. Six individuals’ genetic testing revealed a total of nine pathogenic or likely pathogenic variants, but none were considered explanatory for FIRES. Individuals 9, 10, 14, and 25 had single, heterozygous pathogenic or likely pathogenic variants in autosomal recessive genes identified on epilepsy panels or exome sequencing and were not considered explanatory. Exome sequencing revealed that Individual 2 harbored a heterozygous pathogenic variant in *SCN5A*, associated with autosomal dominant cardiac conduction system dysfunctions and cardiomyopathy, and Individual 24 had a heterozygous pathogenic variant in FLG, associated with autosomal dominant eczema. Mitochondrial genome sequencing revealed that individual 14 had a likely pathogenic variant in MT-TK detected at approximately 4% heteroplasmy in blood and 3% heteroplasmy in brain tissue. Mitochondrial MT-TK variants are most associated with myoclonic epilepsy with ragged-red fibers (MERRF) syndrome at heteroplasmy levels markedly greater than in this individual[25] therefore, it is unlikely that this variant was explanatory for this individual’s clinical features.

We reviewed variants of unknown significance (VOUS) to identify candidate genes, but none were identified. There were 36 total VOUS revealed across all testing modalities. An epilepsy panel revealed a de novo VOUS in ITPR1, associated with autosomal dominant Gillespie syndrome and adult-onset spinocerebellar ataxia, in Individual 9 which did not fit the phenotype for FIRES. Exome sequencing revealed biallelic, compound heterozygous VOUSs, with confirmed inheritance from each parent respectively, in *SPTBN5*. This gene was a candidate gene at the time of this individual’s exome sequencing. While *SPTBN5* has been identified as causative of an autosomal dominant disorder characterized by developmental differences and seizures [26], this gene has not yet been fully validated, and thus the variants found in our patient remain of uncertain clinical significance. Furthermore, given that all known affected individuals with *SPTBN5* had de novo variants, this finding is unlikely to be relevant to the diagnosis of Individual 9. In summary, no genetic etiology was identified for any patient presenting with FIRES.

### Characterization of common genetic etiologies in individuals with SE and RSE suggest at a distinct genetic architecture underlying FIRES

As all genetic testing performed in our FIRES cohort was non-explanatory, we expanded the scope of our study to assess the genetic landscape of a broader group of individuals with SE and RSE, aiming to elucidate how FIRES fits more broadly into the context of SE and RSE. First, we identified 959 individuals with presence of RSE documented in 166,301 time-stamped patient encounters in a broader cohort of 32,112 individuals with childhood epilepsy (Figure 4A). The highest proportion of individuals had onset of RSE during the first three months of life, which contrasts with the distribution of onset in our cohort of individuals with FIRES.

**Figure 4.**
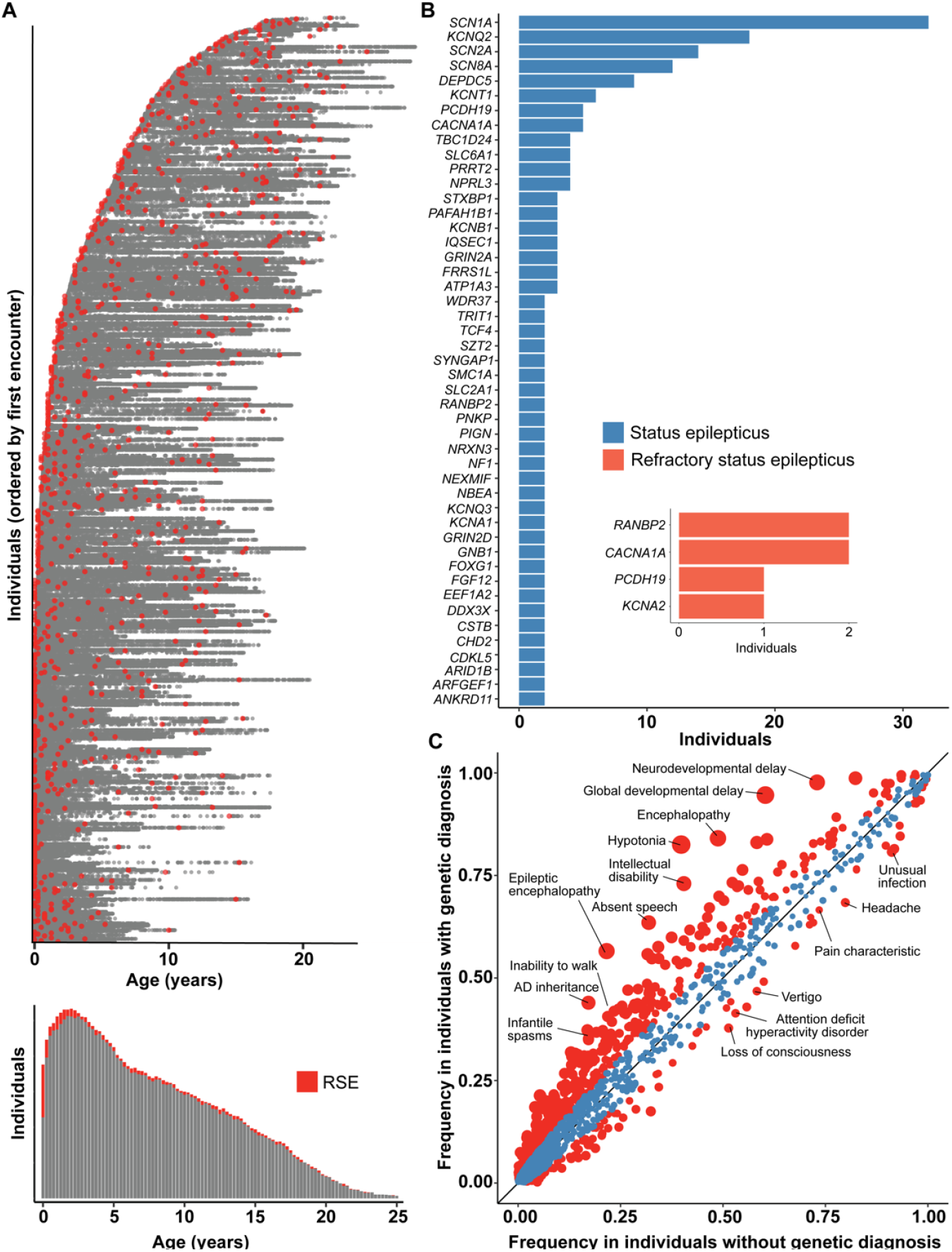
The genetic architecture of status epilepticus and refractory status epilepticus (RSE) differs from FIRES. **(A)** 166,301 time-stamped encounters from the Electronic Medical Records (EMR) across 959 individuals identified with RSE in a broader cohort of 32,112 individuals with childhood epilepsy, showing in red the encounters at which RSE was first documented for each individual. Only the encounter of first RSE onset is captured for each individual, as later encounters with RSE documented could either refer to a new or prior RSE event, and we found that the majority of individuals presenting with RSE have onset within the first three months of life. **(B)** Status epilepticus and RSE in the most common genetic etiologies in a cohort of 1,894 individuals with known or presumed genetic epilepsies. NLP was performed only on patient notes prior to a genetic diagnosis to adjust for bias in clinical impression following a molecular diagnosis. **(C)** Clinical features in 1,158 individuals with status epilepticus stratified by individuals with a genetic diagnosis (n=389) compared to individuals without a genetic diagnosis (n=769), highlighting a difference in overall disease severity between the two subgroups. Red indicates phenotypic features with nominal significance (p<0.05) while size of points indicate -log10(p-value). The landscape of FIRES was distinct from both subgroups, with characteristic severe clinical presentations and no currently identified genetic etiology.

Second, to better understand the genetic landscape of individuals presenting with RSE as well as SE more broadly, we examined our CHOP cohort, narrowing to evaluate only those individuals who had experienced SE or RSE. This yielded a genetic etiology for 36% of cases (Figure 4A). Regarding the occurrence of SE, greater than 80% of individuals carrying certain genetic diagnoses, including *KCNT1, DEPDC5*, and *NPRL3*, had at least one occurrence of SE prior to the genetic diagnosis. For other common genes in our cohort, including *STXBP1, SCN1A, KCNQ2, SCN2A*, fewer than 50% of individuals had at least a one-time presentation with SE prior to genetic diagnosis.

In our CHOP cohort of individuals with SE, we searched for and curated for a smaller subgroup of individuals who presented with RSE as the initial presentation of seizures as a comparative population to individuals with FIRES. After filtering the dataset and excluding individuals following manual review, we identified six individuals with RSE on initial seizure presentation who ultimately were diagnosed with a genetic disorder (Figure 4B). These individuals do meet criteria for NORSE by consensus guidelines. These included one individual with *PCDH19*, two individuals with *CACNA1A*, two twin sisters with homozygous variants in *RANBP2*, and one individual with *KCNA2*. Each of these individuals presented prior to the age of 24 months, and only one presented with RSE after the age of 12 months. Each had genetic testing sent within a week of presentation, except for the individual with PCDH19, for whom data are not available (Supplementary Table 4).

Finally, we performed a phenotypic analysis comparing clinical features such as neurodevelopmental and other epilepsy phenotypes between SE associated with genetic diagnosis (n=389) and idiopathic (non-FIRES) SE (n=769). Individuals with identified genetic etiologies were more likely to have hypotonia (OR 7.13, 95% CI 5.25-9.77), global developmental delay (OR 11.49, 95% CI 7.21-19.24), and epileptic encephalopathy (OR 4.72, 95% CI 3.60-6.21). Individuals without an identified genetic diagnosis had a two-fold higher risk of headache (OR 1.83, 95% CI 1.37-2.44), attention deficit hyperactivity disorder, (OR 1.63, 95% CI 1.24-2.06) and memory impairment (OR 2.09, 95% CI 1.40-3.18) (Figure 4C). This highlights the higher frequency of certain neurological clinical features in individuals with genetic epilepsies.

## Discussion

FIRES is a rare and severe condition characterized by new onset RSE that presents following a febrile illness prior to seizure onset with an unknown pathophysiology and etiology. Previous studies have sought to understand an underlying genetic cause of FIRES. However, thus far findings remain mixed and inconclusive, despite phenotypic commonalities and clinical overlap with other developmental and epileptic encephalopathies. This finding is surprising, given the overall genetic landscape of epilepsy, where yield for genetic testing is 33% [9, 10]. Here, we mapped the landscape of 25 individuals with FIRES, providing an overview of the clinical and treatment histories across 75 cumulative patient-months. We then examined the genetics of broader cohorts of individuals with SE or RSE to better understand how FIRES fits into the conceptual framework of genetic and genetic predispositions in epilepsy.

### Clinical presentation and treatment of children with FIRES

Our cohort consisted of 25 individuals who met clinical criteria for FIRES. The initial clinical symptomatology described in this cohort is in line with other previously published series in children [18, 27]: at onset, children were largely school age, were neurodevelopmentally typical, and exhibited prodromal symptoms prior to onset including confusion, headache, gastrointestinal symptoms, or mild febrile illness/upper respiratory infection. In four individuals, a viral pathogen was identified. Three individuals were diagnosed with an autoimmune condition. All patients had prolonged hospitalizations and a biphasic course characterized by an initial acute catastrophic phase followed by a chronic phase, with refractory epilepsy and some degree of neurological impairment.

Treatment for FIRES was also similar to previously published cohorts [3, 27, 28]. All children received anesthetic infusions during the initial presentation and required anti-seizure medications at the time of discharge. The vast majority of children received steroids and IVIG. Despite the reported efficacy for ketogenic diet [4, 28] only approximately half of these patients received the diet, although the number of individuals with ketogenic diet increased across the years. PLEX was also commonly administered but not as frequently as steroids or IVIG. Notably across years, there was increased use of immuno-modulatory agents including the recombinant version of human IL1RA, anakinra, and the IL-6 pathway antagonist tocilizumab, roughly corresponding to their introduction as a potential efficacious treatment for FIRES in 2016 and 2018, respectively [29, 30].

### Absence of known etiologies in FIRES

First, despite a comprehensive review of genetic testing and review of all variants in our cohort of 25 individuals with FIRES, no genetic etiologies were identified. These negative findings are in agreement with our current understanding of FIRES as described in published literature and through ongoing efforts of etiological discovery [20, 21]. While there are select reports of variants in *SCN1A, PCDH19, POLG, DNM1, KCNT1*, and *SCN2A* linked to FIRES, none of these variants are considered explanatory for the patients’ disease or upon closer review, the clinical presentation does not meet criteria for FIRES [15, 16, 17, 31, 32, 33, 34]. Consequently, despite the high yield for genetic findings in up to 33% of individuals with epilepsy, genetic yield in FIRES is 0% at this time.

The low yield of genetic testing in FIRES may suggest a novel genetic mechanism, polygenic etiology, or alternative etiologies. Inflammatory or autoimmune causes may contribute to the etiology of FIRES, however, given consistently poor outcomes and response to immunomodulatory medications, the latter is unlikely to be the sole explanatory mechanism. It is likely that the underlying etiology is multifactorial and involves a constellation of dysfunctional pathways such as cytokine-mediated inflammation, mitochondrial dysregulation, genetic susceptibility, and environmental exposures. Yet, while it is critical to consider that individuals presenting with RSE may in fact have distinct clinical disorders, the homogeneity of phenotype and consistent absence of genetic findings in FIRES points to a conceptual difference that characterizes FIRES as a singular clinical entity and distinguishes this cohort from both genetic and other forms of idiopathic SE and RSE.

### Conceptual differences between FIRES and epilepsies associated with SE/RSE

While distinct in some features, FIRES is similar to other forms of developmental epileptic encephalopathies such as *STXBP1* and *CKDL5*, in that almost universally all individuals have poor outcomes with a similar phenotypic landscape with cognitive, speech and motor impairment. However, a clinical characteristic that may distinguish FIRES from other forms of RSE is age of onset (Figure 4A). As we found in our broader cohort of 959 individuals with RSE, the highest proportion of individuals with genetic RSE had onset within the first three months of life, highlighting the importance of genes that are variably expressed at different ages and developmental stages. Individuals with FIRES, on the other hand, typically have onset in childhood, with an age of onset ranging from 7.6 months to 18.7 years in our cohort.

To better understand the genetic landscape of SE/RSE more broadly, we aimed to provide an in-depth overview of the landscape in non-FIRES epilepsies associated with SE and RSE. In broadening our cohort to include non-FIRES SE and RSE, we attempted to capture individuals along a gradient of disease severity and onset of seizure presentation, first capturing any individuals prior to genetic diagnosis who were documented to have SE, followed by narrowing our comparison cohort to individuals with RSE, and then only to individuals who had no other seizures prior to the onset of RSE. First, we found that the relative frequency of SE in specific genetic etiologies varies. The underlying architecture of SE is well established, with over 100 genes identified with conditions including inborn errors of metabolism and congenital disorders, structural malformations, mitochondrial disorders, and infantile/childhood onset epileptic encephalopathies, among others [11, 35]. We then narrowed in on individuals without pre-existing epilepsy diagnoses who presented with RSE as their first seizure episode, in order to more closely compare these individuals to children with FIRES. Accordingly, we narrowed in on a much smaller cohort of individuals with RSE who were ultimately diagnosed with genetic conditions and demonstrated that the frequency of genetic diagnoses occurs on a gradient, with fewer and fewer genetic etiologies identified as we narrow our cohort and approach NORSE-like and FIRES-like presentations.

Through comparison with a larger SE/RSE cohort, we demonstrate that there are more recognized genes associated with RSE that require further investigation. As neonatal-onset RSE is clinically distinct from RSE following the neonatal period, we excluded individuals with neonatal onset RSE secondary to variants in genes such as *SCN2A* and *KCNQ2* in assessing genetic etiologies associated with first time presentation of RSE. We subsequently identified four genes that were implicated in individuals with NORSE-like RSE: *CACNA1A, RANBP2, PCDH19*, and *KCNA2*. However, none of these individuals had FIRES, and the lack of substantial evidence in the explanatoriness of these genes in individuals with confirmed FIRES further highlight the complexity of genetic testing and interpretation, underscoring the critical need for ongoing genetic testing in order to generate more evidence for gene and variant validity.

### Comprehensive genetic testing in FIRES

While we have demonstrated an absence of identifiable causative etiologies with our current understanding in our FIRES cohort, the presumed genetic contribution points to the critical need for further elucidation of the underlying genetic landscape to identify pathogenic mechanisms in RSE/SRSE. We show that genetic testing has been heterogeneous throughout the years, particularly with regard to timing of initial test as well as choice of first genetic test. Accordingly, we suggest that genetic testing should be performed as early as possible in the course of the patient’s illness and hospital admission, via Rapid Whole Exome Sequencing with mitochondrial DNA. Although FIRES is distinct from first presentations of genetic SE/RSE, there may be phenotypic commonalities at initial presentation. Thus, finding a genetic diagnosis earlier would likely alter the management course significantly, allowing for more targeted therapies and avoiding unnecessary and potentially harmful immunomodulatory medications. With our current level of genetic understanding, negative genetic testing may aid in narrowing diagnostic differentials, given that individuals with FIRES have been consistently shown not to have a genetic etiology identified on genetic testing. Given the rarity of FIRES, more frequent genetic testing may allow for the discovery of novel, previously unrecognized genes related to seizure susceptibility/epileptogenesis, cellular dysfunction, or immunological dysregulation, that may help in guiding targeted therapies for FIRES and preventing the devastating sequelae. Given the poor yield of alternative genetic testing modalities overall, Whole Exome Sequencing with mitochondrial DNA stands as the most comprehensive, efficient, and cost-effective testing and has been considered the gold standard for diagnostic testing. While we have the capability to perform NGS, this has not been routinely done as demonstrated in our cohort, and even recent consensus guidelines, genetic testing was not recommended as an initial test but only as a later tier of testing [36].

### Limitations

To investigate the overall clinical and genetic landscape of SE and RSE, we leveraged a computational approach based on Natural Language Processing (NLP) for extracting unstructured clinical data. The automatic assessment of clinical text poses challenges in clinical interpretation of large-scale datasets and requires consideration of potential biases. For example, for individuals with a genetic diagnosis, we were limited to assessing clinical features only prior to the diagnosis to prevent “note contamination”, or the bias when phenotypes associated with the genetic diagnosis more broadly is captured in the patient chart and does not necessarily describe or is inaccurate to the specific patient. Furthermore, identification of individuals with FIRES and distinguishing RSE from SE required subsequent manual chart review. In the latter case, we had to review on an individual basis to confirm critical details of case presentations. Nevertheless, we demonstrate that a data-driven method can facilitate the identification of individuals that meet predefined criteria such as FIRES or RSE across a large EMR database and healthcare system that captures dynamic medical care over the years, allowing us to identify 25 individuals who were ultimately determined to meet FIRES criteria and 6 individuals with inaugural RSE secondary to a genetic diagnosis.

Another limitation of our study was the limited analysis of longitudinal clinical data, including the characterization of epilepsy histories and developmental trajectories over time following the initial presentation of FIRES. Furthermore, given the importance of understanding real-world clinical care, the reconstruction of medical treatment following hospital admission would be clinically meaningful. While anesthetic infusions are captured in a standardized framework and level of granularity to the level of a minute timescale in the EMR, data was limited in our cohort due to transfer of care and continuous observation at our clinical center. However, in our study, we provide an objective picture of the heterogeneity in medical treatment using real-world data, including genetic testing in our cohort over the last decade for individuals with FIRES. We point to the importance of an early and comprehensive genetic work-up and the need for future studies to focus on assessment of longitudinal outcomes and trajectories in FIRES, which can be stratified by treatment strategies.

## Conclusion

In our study, we analyzed the phenotypic and genetic landscape of febrile-infection related epilepsy syndrome (FIRES) and introduced a conceptual framework outlining the identification and assessment of clinical histories of 25 individuals with FIRES through a computational approach in the EMR. Comparing FIRES to other epilepsies characterized by SE, we identified a gradient of diagnostic yield and genetic diagnosis and a spectrum of disease severity associated with RSE and NORSE-like presentations (Figure 1, inset). We demonstrated a new paradigm for the consideration of genetic epilepsy, where identifiable genetic etiologies become increasingly rare with the increasing severity of seizure presentation, from SE to RSE, culminating in NORSE, and FIRES. This phenotypic pathway analysis points to a delineation of FIRES from similar conditions associated with explosive onset of epilepsy and RSE and highlights the critical need for future studies investigating underlying etiologies.

## Data Availability

All data produced in the present study are available upon reasonable request to the authors.

## Author Contributions

DD, JX, AK, IH contributed to the writing of the manuscript and figures. AG was responsible for data pulling using systematic computational analysis. DD conducted the manual review of all patients with FIRES and AK, JX conducted the analysis for larger cohorts in this study. KRS, SR and AK were responsible for interpretation of genetic testing from our CHOP and FIRES cohort. PG, MR, NSA were involved in the editing of this manuscript.

## Acknowledgments

We would like to thank the Department of Biomedical and Health Informatics for their continued support.

