## Supplementary Materials for "The genetic spectrum of febrile infection-related epilepsy syndrome (FIRES) and refractory status epilepticus"

**Overview of Supplementary Material**

### 1 Supplementary Table 1. Individuals excluded from FIRES subgroup (n=34)

### 2 Supplementary Table 2. Individuals excluded from FIRES subgroup with NORSE (n=7)

### 3 Supplementary Table 3. Genetic testing in our cohort of 25 individuals with FIRES

### 4 Supplementary Table 4. Individuals with RSE at initial seizure presentation

**Supplementary Table 1. Individuals excluded from FIRES subgroup (n=34)**

| **Diagnoses for individuals excluded from the FIRES cohort** | **Individuals** | **Reason for exclusion** |
| --- | --- | --- |
| Acute necrotizing encephalitis of unknown etiology | 1 | No fever prodrome, history of seizure 2 weeks prior |
| GEFS+ | 1 | History of prior seizures |
| Angelman’s syndrome | 1 | History of prior seizures |
| *SCN2A* | 1 | History of neonatal seizures, epilepsy |
| Acute necrotizing encephalitis with HHV6 virus, *RANBP2* | 1 | Did not have seizures |
| *SCN1A* | 1 | History of prior seizures |
| *PCDH19* | 1 | pre-existing history of seizures, prior to status presentation |
| Idiopathic epilepsy | 4 | History of prior seizures, did not have RSE |
| Pyruvate Carboxylase deficiency | 1 | Metabolic disorder |
| Hypoxic Ischemic Encephalopathy | 2 | Acquired acute symptomatic seizures |
| Epilepsia partialis continua | 1 | History of prior seizures |
| LIG3 mitochondrial encephalopathy | 1 | Did not have RSE |
| MOG encephalitis | 1 | Did not have seizures |
| Febrile status epilepticus | 1 | Fevers at onset of status epilepticus |
| MELAS | 1 | Long standing illness, prior seizures |
| *CACNA1A* | 1 | Febrile with seizure onset |
| FIRES | 8 | Remote history in another country lacking documentation  *One patient with +GAD antibodies, another with history of HSV encephalitis* |
| NORSE* | 6 | Remote history in another country/lacking documentation |

* See Supplementary Table 2 for individuals excluded with NORSE.

**Supplementary Table 2. Individuals excluded from FIRES subgroup with NORSE (n=7)**

| **Individual with NORSE** | **Findings** | **Sufficient documentation for chart review** |
| --- | --- | --- |
| 1 | *PCDH19*, recent vaccination for flu ; NORSE | No; consultation without outside records |
| 2 | NORSE | No; remote history in outside country without documentation |
| 3 | NORSE | No; Telephone consult at CHOP without outside records |
| 4 | NORSE/MOG encephalitis | Yes |
| 5 | NORSE | Yes |
| 6 | NORSE | Yes |

There were 3 individuals with NORSE with sufficient clinical information, therefore further analyses and clinical data on NORSE were excluded from this study.

**Supplementary Table 3. Genetic testing in our cohort of 25 individuals with FIRES**

| Participant | Year diagnosed with FIRES (age, years)* | Single Gene | Karyotype | SNP Microarray | Epilepsy Gene Panel | Exome | Mitochondrial | Other |
| --- | --- | --- | --- | --- | --- | --- | --- | --- |
| 1 | 2010 (5-10 years) |  |  |  |  |  |  |  |
| 2 | 2012 (5-10 years) |  |  |  |  |  |  |  |
| 3 | 2012 (5-10 years) |  |  | Normal result |  |  |  |  |
| 4 | 2013 (10-15 years) | *SCN1A* sequencing report N/A for review |  |  | *• GAMT,* c.581T>C p.(V194A), heterozygous, inheritance unknown, VOUS |  |  |  |
| 5 | 2013 (5-10 years) |  |  | Normal result | *• NRXN1,* c.2627C>T p.(A876V), heterozygous, inheritance unknown, VOUS  *• ZEB2,* c.808-4A>G p.? heterozygous, inheritance unknown, VOUS |  |  |  |
| 6 | 2013 (15-20 years) |  | Normal result | Normal result |  |  |  |  |
| 7 | 2013 (10-15 years) | *TWIST* sequencing: Negative | Normal result |  | Negative |  |  |  |
| 8 | 2014 (0-5 years) |  |  |  |  | *• SCN5A*, c.5477G>A p.(R1826H), heterozygous, inheritance unknown, pathogenic | *• HSPD1*, c.425G>A p.(A142L), heterozygous, VOUS  *• DARS2,* c.228-20dupT, IVS2-20dupT, heterozygous, VOUS | Maturity Onset Diabetes of the Young (MODY) Panel:  *• BLK,* c.26C>T p.(P9L), heterozygous, inheritance unknown, VOUS  WES reanalysis: no additional variants |
| 9 | 2014 (5-10 years) | *SCN1A* sequencing: Negative |  | Normal result |  | *• ITPR1,* c.187A>G p.(M63V), heterozygous, de novo, VOUS • *BTD,* c.1330G>C p.(D444H), heterozygous, maternally inherited pathogenic  *•* *CFTR,* c.1521_1523delCTT p.(F508del), heterozygous, maternally inherited, pathogenic  *•* *COL18A1,* c.2824_2840delGGCCCCCCAGGCCCCCC, heterozygous, inheritance unknown, pathogenic  *•* *POLRIC,* c.796delG heterozygous, paternally inherited, pathogenic | Negative |  |
| 10 | 2014 (0-5 years) |  |  | Normal result | *•* *PNKP,* c.1123G>T p.(G375W), heterozygous, inheritance unknown, likely pathogenic | Duo reanalysis  *•* *CPA6,* c.919G>A p.(A307T), heterozygous, inheritance unknown, VOUS  *•* *PNKP,* c.1123G>T p.(G375W), heterozygous, inheritance unknown, pathogenic | Negative |  |
| 11 | 2014 (5-10 years) | *POLG* sequencing: Negative | Normal result | Normal result | N/A for review |  |  |  |
| 12 | 2016 (0-5 years) |  |  |  | *•* *CNTN2*, c.2735C>T p.(P912L), heterozygous, inheritance unknown, VOUS |  |  | Humoral Dysfunction Panel:  *•* *LRBA*, c.6695T>C p.(I2232T), heterozygous, inheritance unknown, VOUS |
| 13 | 2016 (10-15 years) |  |  |  | Negative | N/A for review |  |  |
| 14 | 2016 (5-10 years) |  | Normal result |  | Negative | Negative | *•* MT-TK m.8342 G>A p.? approximately 3% heteroplasmy, likely pathogenic variant (extracted from brain tissue, approximately 4% heteroplasmy in the blood) |  |
| 15 | 2017 (0-5 years) | *CPT2* sequencing: Negative |  | Normal  result | Negative | *•* *CPA6,* c.932G>A p.(R311Q), heterozygous, maternally inherited, VOUS | Negative |  |
| 16 | 2018 (5-10 years) |  | Normal result |  |  | Negative | *•* *MT-ND5* m.13154T>C p.(I273T), approximately 16%, maternally inherited, VOUS |  |
| 17 | 2018 (0-5 years) |  |  |  | Negative | Negative | *•* *ADCK4*, c.1336G>C p.(G446R), heterozygous, inheritance unknown, VOUS  *•* *MRPS22*, c.766C>T p.(R256C), heterozygous, inheritance unknown, VOUS  *•* *TYMP*, c.1207G>A p.(G403L), heterozygous, inheritance unknown, VOUS | WES Reanalysis: negative |
| 18 | 2018 (5-10 years) | *SCN1A* sequencing: N/A for review    *POLG* - sequencing: N/A for review |  |  | *•* *PNPO,* c.527C>G p.(S176C), heterozygous, VOUS |  |  |  |
| 19 | 2019 (5-10 years) | *HLA-B*: Negative | Normal result | *•* 1.49Mb interstitial duplication within chromosome Xq23, inheritance unknown, VOUS |  | *•* *GRIN2D*, c.250G>C p.(V84L), heterozygous, maternally inherited, VOUS  *•* *SPTBN5,* c.2366G>A p.(R789G), heterozygous, maternally inherited, VOUS  *•* *SPTBN5,* c.749G>T p.(G250V), heterozygous, paternally inherited, VOUS |  | Fragile X: Negative |
| 20 | 2019 (0-5 years) |  |  | Normal result | *•* *JMJD1C* c.3933T>C (silent mutation), het, VOUS |  |  |  |
| 21 | 2019 (5-10 years) |  |  |  |  | *•* ATP1A3, c. 719T>C p.(V240A), heterozygous, paternally inherited, VOUS | Negative |  |
| 22 | 2020 (15-20 years) |  |  |  | N/A for review | *•* *ADAM17,* c.1894G>A p.(V632I), heterozygous, inheritance unknown, VOUS  *•* *ADAM17,* c.178C>A p.(L60I), heterozygous, inheritance unknown, VOUS  *•* *CR2,* c.2518C>G p.(R840G), heterozygous, inheritance unknown, VOUS  *•* *NTRK1,* c.1643G>A p.(R548Q), heterozygous, inheritance unknown, VOUS | Negative |  |
| 23 | 2020 (5-10 years) |  | N/A for review |  | Negative | *•* *LPA,* c.2462C>T p.(P821L), heterozygous, paternally inherited, VOUS | Negative |  |
| 24 | 2020 (5-10 years) |  |  |  | *•* *DIAPH1,* c.2158C>T p.(L720F), heterozygous, paternally inherited, VOUS  *•* *RANBP2,* c.1320A>T p.(L450F) heterozygous, maternally inherited, VOUS | *•* *FLG,* c.8911A>T p.(R2971*), heterozygous, paternally inherited, Pathogenic | Negative |  |
| 25 | 2021 (15-20 years) | *HBB* sequencing:  *•* *HBB,* c.92+5G>C, Intronic, heterozygous, inheritance unknown, VOUS  *HBA1:* Negative  *HBA2:* Negative | Normal result | Normal result | *•* *CACNA2D2*, c.227G>A p.(R76H), heterozygous, inheritance unknown, VOUS  *•* *CNTNAP2,* c.14C>T p.(P5L), heterozygous, inheritance unknown, VOUS  *•* *GTPBP3*, c.1255G>A p.(G419S), heterozygous, inheritance unknown, VOUS  *•* *PACS2*, c.2597T>C p.(V866A), heterozygous, inheritance unknown, VOUS  *•* *PEX10*, c.845T>G p. (L282W), heterozygous, inheritance unknown, VOUS  *•* *RELN*, c.576A>G (Silent), heterozygous, inheritance unknown, VOUS | *•* *DUOX2* c.2895_2898del p.(F966Sfs*29), heterozygous, inheritance unknown, Likely Pathogenic  *•* *SLC9A7,* c.1985T>C p.(L662P), hemizygous, maternal inheritance, VOUS |  |  |

* Ages are provided in ranges of 5 years.

**Supplementary Table 4. Individuals with RSE identified from CHOP cohort**

| **Case** | **Age* (Sex)** | **Clinical presentation** | **Gene and variant** | **Outcome** |
| --- | --- | --- | --- | --- |
| **1** | 20-25 m (F) | Refractory focal status epilepticus in the setting of metapneumovirus | *CACNA1A* p.Leu618Ser | Well-controlled epilepsy and mild developmental delay |
| **2** | 0-5 m (M) | Focal status epilepticus that was initially responsive to one dose of lorazepam, however within a couple hours his seizures recurred, requiring escalating doses of medication | *CACNA1A* p.Val1396Met | Refractory epilepsy, moderate global developmental delay, autism, ataxia, and hemiplegic migraine |
| **3** | 10-15 m (F) | Twins born from consanguinous parents. Both presented with refractory status epilepticus in the setting of a febrile illness. Both had acute brain injury with restricted diffusion and signal abnormalities in the brainstem and deep gray regions in the brain consistent with acute necrotizing encephalitis | *RANBP2*  p.Asp2631Ala | Unknown |
| **4** | 5m-10 m (F) |  | *RANBP2*  p.Asp2631Ala | Developmental delay |
| **5** | 0-5 m  (F) | Refractory clinical SE. Received several doses of anti-seizure medications, then was found to be in subclinical status epilepticus upon initiating EEG | *KCNA2*  p.Pro405Leu | Neurotypical at last follow-up at 7m |
| **6** | 5-10 m (F) | RSE in the setting of fever after vaccine administration | *PCDH19*  p.Thr404Ile | Unknown |

* Ages are provided in ranges of 5 months.
